# Effects of mixed metal exposures on MRI diffusion features in the medial temporal lobe

**DOI:** 10.1101/2023.07.18.23292828

**Authors:** Eun-Young Lee, Juhee Kim, Janina Manzieri Prado-Rico, Guangwei Du, Mechelle M. Lewis, Lan Kong, Jeff D. Yanosky, Paul Eslinger, Byoung-Gwon Kim, Young-Seoub Hong, Richard B. Mailman, Xuemei Huang

## Abstract

**Background:** Environmental exposure to metal mixtures is common and may be associated with increased risk for neurodegenerative disorders including Alzheimer’s disease

**Objective:** This study examined associations of mixed metal exposures with medial temporal lobe (MTL) MRI structural metrics and neuropsychological performance.

**Methods:** Metal exposure history, whole blood metal, and neuropsychological tests were obtained from subjects with/without a history of mixed metal exposure from welding fumes (42 exposed subjects; 31 controls). MTL structures (hippocampus, entorhinal and parahippocampal cortices) were assessed by morphologic (volume, cortical thickness) and diffusion tensor imaging [mean (MD), axial (AD), radial diffusivity (RD), and fractional anisotropy (FA)] metrics. In exposed subjects, correlation, multiple linear, Bayesian kernel machine regression, and mediation analyses were employed to examine effects of single- or mixed-metal predictor(s) and their interactions on MTL structural and neuropsychological metrics; and on the path from metal exposure to neuropsychological consequences.

**Results:** Compared to controls, exposed subjects had higher blood Cu, Fe, K, Mn, Pb, Se, and Zn levels (p’s<0.026) and poorer performance in processing/psychomotor speed, executive, and visuospatial domains (p’s<0.046). Exposed subjects displayed higher MD, AD, and RD in all MTL ROIs (p’s<0.040) and lower FA in entorhinal and parahippocampal cortices (p’s<0.033), but not morphological differences. Long-term mixed-metal exposure history indirectly predicted lower processing speed performance via lower parahippocampal FA (p=0.023). Higher whole blood Mn and Cu predicted higher entorhinal diffusivity (p’s<0.043) and lower *Delayed Story Recall* performance (p=0.007) without overall metal mixture or interaction effects.

**Discussion:** Mixed metal exposure predicted MTL structural and neuropsychological features that are similar to Alzheimer’s disease at-risk populations. These data warrant follow-up as they may illuminate the path for environmental exposure to Alzheimer’s disease-related health outcomes.

**Highlights:**

- Mixed metal exposed subjects through welding fumes had higher blood Cu, Fe, K, Mn, Pb, Se, and Zn levels than controls.
- Exposed subjects had higher diffusion tensor imaging (DTI) mean (MD), axial (AD), radial (RD) diffusivity values in all medial temporal lobe (MTL) regions of interest (ROI) (hippocampus, entorhinal and parahippocampal cortices) and lower fractional anisotropy (FA) in the entorhinal and parahippocampal cortices without significant morphologic differences.
- Long-term mixed metal exposure history predicted altered MTL DTI metrics (lower parahippocampal FA and higher hippocampal RD).
- Across correlation, multiple linear, and Bayesian kernel machine regression analyses, higher whole blood Mn and Cu levels predicted higher entorhinal diffusivity values and lower *Delayed Story Recall* performance, features resembling Alzheimer’s disease at-risk populations.
- MTL DTI metrics mediate, at least partially, the effects of metal exposure on cognitive performance.

## Introduction

There is a growing concern about health consequences of environmental exposure to toxic metals found in ambient air pollution (e.g., vehicle emissions), contaminated land, and occupation-related activities (Crous-Bou et al., 2020; Delgado-Saborit et al., 2021; Lee et al., 2018; Swartjes, 2015). Even essential metals (e.g., Al, Cu, Fe, Mn) can be neurotoxic in lower or higher dosage than its equilibrium status (Teleanu et al., 2018; Zhou et al., 2018), whereas non-essential metals (e.g. Cd, Hg, Pb) can be neurotoxic at low exposure levels (Sanders et al., 2009a). These metals can cross the blood-brain barrier, accumulate in specific brain regions, affect cellular functions, and may contribute to neurodegenerative processes (Banerjee et al., 2014; Liu et al., 2010; Miu and Benga, 2006).

Alzheimer’s disease is the most common age-related neurodegenerative disorder, comprising about 50-70% of dementia cases (Brookmeyer et al., 2011). It is characterized pathologically by accumulation of β-amyloid plaques and tau tangles in the brain, with the most prominent neuronal damage noted in the hippocampus (Li et al., 2019a; Sorensen et al., 2016). Although the major and early disease-related behavioral deficits entail learning/memory problems (Backman et al., 2005), the disease also affects executive, attentional, and language functions (Galton et al., 2000). The clinical diagnosis of dementia is based on a substantial reduction from previous performance levels in more than one cognitive domain that sufficiently interferes with independent daily living (Arvanitakis et al., 2019). At Alzheimer’s diagnosis, hippocampal atrophy (representing significant neuronal loss) is usually apparent (Halliday, 2017). The only disease modifying FDA-approved drugs (e.g., lecanemab) have modest effects on disease progression (Bateman et al., 2022; van Dyck et al., 2023), underscoring the importance of identifying modifiable causes of dementia.

Preclinical studies have shown that metal exposure (e.g., Cu, Mn, and Fe) can lead to Alzheimer’s-like Aβ production in brain and cognitive impairment (Guilarte, 2010; Kitazawa et al., 2016; Pal et al., 2013; Schneider et al., 2013; Schroder et al., 2013; Sobolewski et al., 2022), Epidemiological studies also have reported higher plasma metal levels (e.g., Al, Cd, Cu, Fe, Hg, Pb, and Se) in populations with Alzheimer’s disease and related disorders (Basun et al., 1991; Gerhardsson et al., 2008; Hare et al., 2016; Park et al., 2014; Vural et al., 2010; Xu et al., 2018). Despite the basic science and epidemiological data, linking chronic life-long metal exposure to age-related neurodegenerative diseases in humans, individually or in subgroups, remains challenging. This is due to the difficulty in ascertaining exposure accurately (either distant in time or cumulative, or both), the long-latency of the diseases, and the fact that many early disease-related changes may be subtle and resemble normal aging. Moreover, some metals share exposure sources and pathways through same receptors and/or transporters that can generate effects synergistic or antagonistic to each other (Bauer et al., 2020). Most preclinical and epidemiological studies so far, however, have paid little attention to the mixture or interaction effects of co-exposed metals on neurodegenerative processes.

As the result of decades of research, a number of fluid-based biomarkers (e.g., Aβ and tau in blood and CSF) have promise of being able to reflect Alzheimer’s-related disease process in clinical practice (Bjorkli et al., 2020; Visser et al., 2022; Xie et al., 2022). These biofluid based biomarkers, however, do not yield brain region-specific information and/or underlying mechanistic data about how some brain regions are more vulnerable to disease processes. Radioligand-based neuroimaging [e.g., β-amyloid positron emission tomography (PET)] provides brain-region specific information, and is used in clinical practice, but it requires radiation exposure and is not conducive to studying populations-at-risk in the community. More importantly, radioligand based neuroimaging can only report on one target at a time (Hatt et al., 2017; Vaquero and Kinahan, 2015), limiting its utility to examine multidimensional features related to neurodegeneration.

Promisingly, a growing number of brain MRI techniques can serve as a useful non-invasive biomarker to gauge region-specific neurodegenerative processes, including Alzheimer’s disease-related early changes (Kress et al., 2023; Shukla et al., 2023; Xie et al., 2023). Recent studies suggested that a variety of brain structures (*e.g.*, fornix, cingulum, entorhinal and parahippocampal cortices, posterior cingulate, and precuneus) may exhibit changes early in the Alzheimer’s disease process (Berron et al., 2020; Echavarri et al., 2011; Lacalle-Aurioles and Iturria-Medina, 2023). Among these brain regions, medial temporal lobe (MTL) memory areas that include the hippocampus and entorhinal and parahippocampal cortices may play a key role in Alzheimer’s disease-related cognitive decline, particularly affecting learning/memory (Eichenbaum and Lipton, 2008; Sugar and Moser, 2019). Previous MRI studies reported MTL morphometric (*e.g.*, atrophy, shape) and diffusivity (reflecting microstructural integrity) alterations in both early-stage Alzheimer’s disease and in at-risk populations (*e.g.*, MCI; mild cognitive impairment) that subsequently converted to Alzheimer’s disease (Gerardin et al., 2009; Lancaster et al., 2016; Leandrou et al., 2018). Others have suggested that entorhinal, rather than hippocampal, areas may better predict future Alzheimer’s disease conversion (Killiany et al., 2002; Leandrou et al., 2018).

In the current study, we examined the effects of mixed metal exposures on MRI structural features of MTL structures and cognition in welders using Bayesian kernel machine regression (BKMR), a statistical method that can flexibly model non-linear effects of multiple metal mixtures and their interactions on health outcomes (e.g., MRI/cognition). Welders can serve as an epitype human population for translational research of metal mixture exposure-related neurotoxicity because welding fumes contain a variety of both essential (e.g., Cu, Fe, Mn) as well as non-essential (e.g., Pb, Ni, Cd, Cr) metals (Pajarillo et al., 2021; Sanders et al., 2009b; Yarjanli et al., 2017). Our overall hypothesis was that exposure to metal mixture via welding fumes is associated with MTL structural and cognitive changes as seen in Alzheimer’s disease at-risk populations. We tested three specific hypotheses: **H1**) Exposed subjects (welders) will have higher medial (MD), axial (AD), radial (RD) diffusivity, and lower fractional anisotropy (FA) values in MTL structures than controls; **H2**) Exposed subjects (welders) will have lower performance on processing/psychomotor speed, executive, and learning/memory tasks than controls; and **H3**) In exposed subjects (welders), multiple metal exposures will predict MTL structural and cognitive metrics as separate single but also as mixture. Lastly, we explored potential mediation effects of mixed-metal exposure on neuropsychological performance via MTL diffusion features.

## Methods

### Study subjects

Eighty subjects were recruited from central Pennsylvania (USA). Exposed subjects were welders who had welded at any point in their lifetime, whereas controls had no such history. Enrollment occurred between 2011-2013 and overlapped such that both groups were recruited equally over this timeframe. All subjects denied past Parkinson’s disease or Alzheimer’s diseases diagnoses or other neurological or neuropsychiatric disorders. All subjects were male, with Movement Disorder Society Unified Parkinson’s disease Rating Scale (Goetz et al., 2008) motor exam sub-scores (UPDRS-III) < 15, Mini-Mental Status Examination (MMSE) scores >24, and Montreal Cognitive Assessment (MoCA) >19. Forty-two welders and 31 controls completed brain MRI acquisition with good quality images (five welders and two controls were excluded). Written informed consent was obtained in accordance with the Declaration of Helsinki and approved by the Penn State Hershey Internal Review Board.

### Ascertainment of welding fume exposure and metal blood levels

We utilized established metal exposure questions(Lee et al., 2015) to estimate the following metrics. 1) Recent metal exposure occurring in the 90 days prior to the study visit including hours welding [*HrsW_90_* = (weeks worked) * (h/week) * (fraction of time worked related directly to welding) and *E_90_* (an estimate of the cumulative metal exposure via welding)]. 2) The lifetime metal exposure including welding years [(*YrsW*=years spent welding during the subjects’ life) and *ELT* (an estimate of cumulative metal exposure via welding fumes over the individual’s life)] (Lee et al., 2015).

We obtained whole blood the morning (ca. 0800 h) of the same day that cognitive tests and brain MRI were performed. The whole blood samples were analyzed by Inductively Coupled Plasma Mass Spectrometry (ICP-MS) for trace minerals including Ca, Cr, Cu, Fe, K, Mg, Mn, Na, Pb, Se, Sr, and Zn. Digestion was performed by microwave methods using the Discovery SPD digestion unit (CEM, Matthews, North Carolina). After digestion, the samples were analyzed for trace minerals using the Thermo (Bremen, Germany) Element 2 SF-ICP-MS equipped with a concentric glass nebulizer and Peltier-cooled glass cyclonic spray chamber. Bulk mineral concentrations were determined by ICP-OES (Optical Emission Spectrometry) analysis on the Thermo iCAP equipped with a polypropylene cyclonic spray chamber at the University of North Carolina, Chapel Hill, NC, USA (Lee et al., 2015).

### Neuropsychological tests (NPTs) and scores

Six neurobehavioral domains were examined: (1) processing/psychomotor speed; (2) executive function; (3) language; (4) learning/memory; (5) visuospatial processing; and (6) attention/working memory. Subtests from the *Repeatable Battery for the Assessment of Neuropsychological Status (RBANS)(Randolph et al., 1998)* were used to assess language, visuospatial processing, and learning and memory. Processing/psychomotor speed, executive function, and attention/working memory domains were evaluated using subtests from: the *Wechsler Adult Intelligence Scale-Third Edition* (*WAIS-III)* (Wechsler, 1997); *Trail Making Tests (A and B)* (Tombaugh, 2004); *Stroop Test* (Golden, 1978), *Delis-Kaplan Executive Function System (D-KEFS)* (Delis et al., 2001); and *Wechsler Memory Scale-Third Edition* (*WMS-III*) (Wechsler, 1998). Tests were administered by an experienced examiner. Specific domains and tests follow.

#### Processing/psychomotor speed

During the *Symbol Search* subtest, subjects were presented with a set of symbols and asked to report if the provided set contained a particular symbol. In the *Stroop-Word* test (Golden, 1978), color words (e.g., “RED,” “GREEN,” and “BLUE”) were presented randomly in black ink. Subjects were asked to read the words as quickly as possible. In the *Stroop-Color* test (Golden, 1978), each item was written as “XXXX” printed either in red, green, or blue. Subjects were asked to name the colors as quickly as possible. During the *Trail Making-A* test, subjects were required to connect numbers from 1 to 25 consecutively as quickly as possible.

#### Executive function

In the *Symbol-Digit Coding* subtest, subjects performed a timed task to match symbols to designated numbers. During the *Stroop-Color-Word* subtest, the words “RED,” “GREEN,” and “BLUE” were printed in non-corresponding colors (e.g., the word “RED” printed in green ink). Subjects were asked to name the ink color of the printed words. In the *Trail Making-B*, subjects were required to connect numbers (from 1 to 13) and letters (from A to L) consecutively in an alternating sequence (e.g., 1-A-2-B etc.). During the *Phonemic Fluency* subtest from *D-KEFS*, subjects generated words beginning with a particular letter (e.g., “F”) within one minute.

#### Language

In the *Picture Naming* test, subjects were asked to name 10 familiar objects. During the *Semantic Fluency* test, subjects generated as many words within a restricted category (fruits/vegetables or zoo animals) as possible within one minute.

#### Learning/memory

In the *List Learning* test, a non-organized list of 10 words was presented orally and subjects immediately recalled the word list. Four learning trials were performed where subjects were encouraged to recall the words in any order. During the *Delayed List Recall* test, subjects recalled the same word list after ca. 20-min delay. For the *Immediate Story Recall* test, subjects were presented with a short prose passage and immediately reproduced the story they heard. In the *Delayed Story Recall* test, subjects recalled the story after ca. 20-min delay. For the *Delayed Figure Recall* test, subjects reproduced a figure after a ca. 20-min delay.

#### Visuospatial processing

This was evaluated with *Line Orientation* and *Figure Copy* subtests from the *RBANS.* In the *Line Orientation test,* subjects were presented with a pair of angled lines and asked to match them to two numbered lines on the display. During the *Figure Copy* subtest, subjects copied a complex figure.

#### Attention/working memory

In the *Digit Span* test, subjects orally were presented with a series of increasing digits, and asked to recite the digits in the presented or reversed order. In the *Spatial Span* test, a three-dimensional board with ten blocks was used and a series of increasing blocks were presented in a specific, predetermined pattern. Subjects were asked to reproduce the same pattern by pointing to the blocks in forward or reverse order. In the *Letter-Number Sequencing* test, subjects orally were presented with mixed sequences of digits and letters and asked to reproduce the digits in ascending order, followed by the letters in alphabetical order.

Individual raw neuropsychological scores were converted to age-adjusted norm scores (e.g., T-scores or scaled scores). To unify these scores, the normative scores then were converted to z-scores [(individual norm-based– mean norm) / (standard deviation of the norm)]. Z-scores for the *Stroop (Stroop-Word, Stroop-Color, and Stroop-Color-Word), Trail Making*-*A* and -*B, Phonemic Fluency, and Digit Span* tests were calculated based on age- and education-adjusted normed scores (Gladsjo et al., 1999; Golden, 1978; Monaco et al., 2013; Ruff and Parker, 1993; Tombaugh, 2004). *Phonemic Fluency* subtest scores additionally were adjusted for race (Gladsjo et al., 1999).

### MRI acquisition and processing

All MR images were acquired using a Siemens 3 T scanner (Magnetom Trio, Erlangen, Germany) with an 8-channel head coil. First, high-resolution T1-weighted (T1W) and T2-weighted (T2W) images were acquired for anatomical segmentation. For T1W images, MPRAGE sequences with Repetition Time (TR)/Echo Time (TE)=1540/2.3 ms, FoV/matrix=256×256/256×256 mm, slice thickness=1 mm, slice number=176 (with no gap), and voxel spacing 1×1×1 mm were used. T2W images were obtained using fast-spin-echo sequences with TR/TE=2500/316 ms and the same spatial resolution as the T1W images. For R1, TR/TE=15/1.45 ms, flip angles=4/25°, FoV/matrix=250×250/160×160 mm, slice thickness=1 mm, slice number=192, and voxel spacing=1.56×1.56×1 mm were used. R2* images were acquired using five TEs ranging from 8-40 ms with an interval of 8 ms, TR=51 ms, flip angle=15°, FoV/matrix=230×230/256×256 mm, slice thickness=1.6 mm, and slice number=88 were used. For DTI, TR/TE=8300/82 ms, b value=1000 s/mm^2^, diffusion gradient directions=42 and 7 b=0 scans, FoV/matrix=256×256/128×128 mm, slice thickness=2 mm, and slice number=65 were used.

#### Regions of Interest

Medial temporal regions that previously had been reported as Alzheimer’s disease-related early changes (hippocampus, and entorhinal and parahippocampal cortices; Figure 1) were selected as regions-of-interest (ROIs). They were defined for each subject using Freesurfer (Anonymous, 2013). Segmentation quality then was confirmed visually by a reviewer blinded to group assignment.

**Figure 1.**
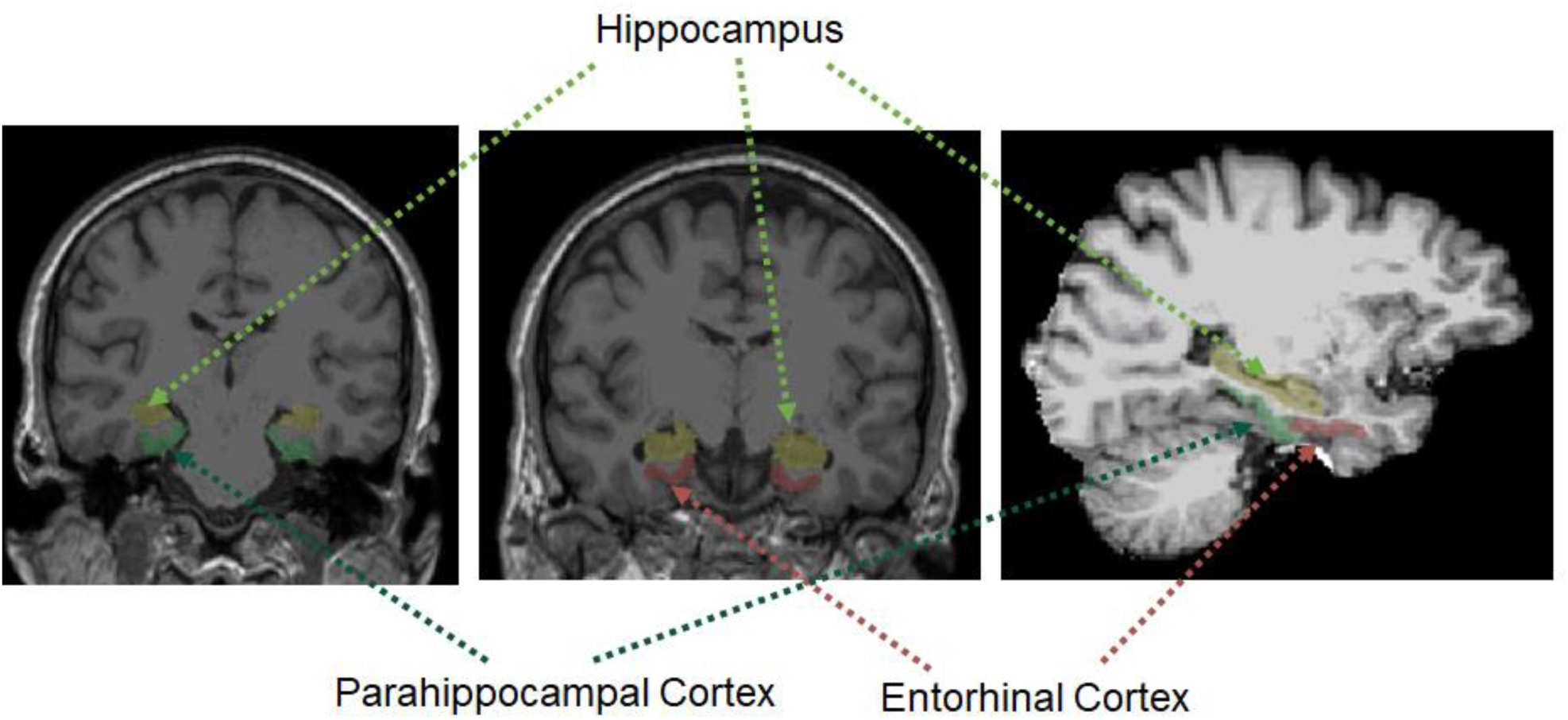
Automatically segmented regions of interest [hippocampus, entorhinal and parahippocampal cortices] on T1-weighted MPRAGE images.

#### Morphologic Metrics

Hippocampal segmentation for volume estimation and cortical parcellation for thickness calculation was performed with the Freesurfer image analysis suite (http://surfer.nmr.mgh.harvard.edu/). Processing included motion correction, removal of non-brain tissue using a hybrid watershed/surface deformation procedure (Segonne et al., 2004), and automated Talairach transformation. Within temporal lobe, deep gray matter volumetric structures including the hippocampus then were segmented and cortical gray matter structures including entorhinal and parahippocampal cortices were parcellated (Fischl et al., 2002; Fischl et al., 2004).

#### Diffusion Metrics

DTI quality control and tensor reconstruction were performed using DTIPrep(Oguz et al., 2014) that first checks diffusion images for appropriate quality by calculating the inter-slice and inter-image intra-class correlation, and then corrects for the distortions induced by eddy currents and head motion. DTI maps then were estimated via weighted least squares. The DTI maps were co-registered onto T1/T2W images using ANTS, and the transformation matrix was applied inversely to bring the hippocampal region to the DTI maps. Four DTI indices [fractional anisotropy (FA), AD (axial diffusivity), RD (radial diffusivity), and MD (mean diffusivity)] were calculated out of three diffusivity eigenvalues (λ1, λ2, λ3) (Le Bihan et al., 2001). FA is a weighted average of pairwise differences of the three eigenvalues and may represent the degree of diffusion anisotropy. AD is the largest eigenvalue (λ1) and RD is an average of the remaining two eigenvalues, both of which may indicate the orientation of the diffusion. MD is an average of the three eigenvalues, providing the overall diffusion magnitude (Le Bihan et al., 2001).

### Statistical analysis

Group comparisons of demographic data were conducted using one-way analysis of variance (ANOVA). For group comparisons of metal exposure history and whole blood metal levels, multivariate analysis of variance (MANOVA) was used to control intercorrelations among dependent variables. Whole blood metal levels were log-transformed to attenuate the impact of potential extreme values. Group comparisons of neuropsychological and neuroimaging metrics were conducted using multivariate analysis of covariance (MANCOVA). Adjusted neuropsychological z-scores were used for the group analyses, with adjustment for education level if it was not adjusted individually. For cortical thickness and DTI measures, analyses included adjustment for age and education level. When comparing hippocampal volume, total intracranial volume (TIV) also was used as a covariate.

Multiple statistical analyses were conducted to understand the relationship between multiple metal exposure dose-health outcome (brain structural and neuropsychological metrics) response relationship in exposed subjects. **<u>First,</u>** Pearson partial correlation analyses were used with adjustment for potential confounders. For the associations of neuropsychological and MRI metrics with metal exposure history, age and education level were used as covariates. For the associations of neuropsychological and MRI metrics with individual blood metal levels, other blood metal levels were adjusted in addition to age and education. For the associations between neuropsychological and MRI metrics, age and education levels were used as covariates. **<u>Second,</u>** multiple linear regression analyses were conducted. For these analyses, neuropsychological and MRI metrics were treated as outcome variables, whereas metal exposure history and blood metal levels were treated as predictors with adjustment for age and education. For neuropsychological scores, multiple linear regression analyses additionally were conducted while treating MRI metrics as predictors with adjustments of age and education. **<u>Third,</u>** BKMR analyses(Bobb et al., 2015) were conducted to provide flexible modeling of non-linear and non-additive associations between exposure predictors of multiple metals and outcome variables (MRI/cognition). We used a Markov Chain Monte Carlo (MCMC) algorithm while treating covariates as linear. These analyses allow testing of the overall metal mixture and its pairwise interaction effects in addition to single exposure variable effects. **<u>Lastly,</u>** causal mediation analyses (Ding and VanderWeele, 2016; Valeri and Vanderweele, 2013; VanderWeele, 2015; Zhang et al., 2016) were conducted to test whether neuropsychological metrics were linked to metal exposure measurements either directly, or indirectly via MTL structural metrics, with age and education level included as covariates (Figure 2). To test mediation effects of blood metal on neuropsychological metrics, other blood metals additionally were used as covariates. A statistical significance level of α=0.05 was used. Correlation analyses of neuropsychological and MRI metrics with metal exposure measures were corrected for multiple comparisons using the Stepdown Bonferroni method (Holm, 1979) to control the familywise error rate (*FWER*) at *p*=0.05. We report uncorrected raw *p*-values but indicate significant results after *FWER*-correction. SAS 9.4 or R was used for statistical analyses.

**Figure 2.**
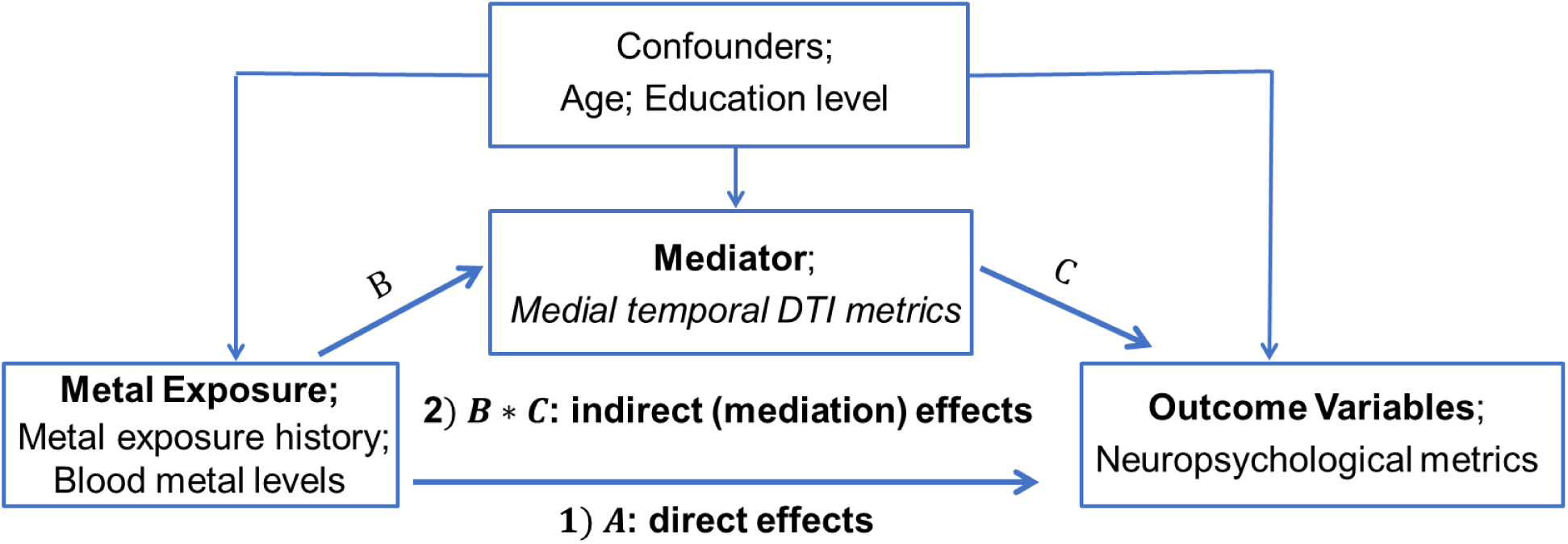
Mediation models to test effects of exposure (metal exposure history; blood metal levels) on neuropsychological metrics while medial temporal DTI metrics serve as mediators after controlling for potential confounders (age; education level; blood levels of other metals): 1) A indicates direct effects of exposure on neurobehavioral metrics that are not mediated by medial temporal DTI metrics; 2) B*C indicates mediation effects of exposure on neurobehavioral metrics that are mediated by medial temporal DTI metrics.

## Results

### Demographics and exposure features of study participants

The exposed subjects (welders) were older (p=0.048) and had lower education years (p<0.001) than controls. There was no significant group difference in UPDRS-III or MoCA scores (p=0.326 and p=0.797, respectively; Table 1a). Exposed subjects displayed significantly higher exposure metrics (p’s<0.001) and whole blood metal levels of Cu, Fe, K, Mn, Pb, Se, and Zn (p’s<0.026; Table 1b).

**Table 1.** Summary statistics for demographics (a) and overall exposure measures (b) for controls and exposed subjects.

|  | Controls<br>(N=31) | Exposed<br>subjects<br>(N=42) | Raw<br>p-values |
| --- | --- | --- | --- |
| ***a. Demographics*** |  |  |  |
| Age (y) | 43.6 ± 11.4 | 48.9 ± 10.7 | 0.048 |
| Education (y) | 16.2 ± 2.2 | 12.9 ± 1.6 | < 0.001 |
| MoCA | 26.2 ± 2.5 | 26.3 ± 2.1 | 0.797 |
| UPDRS-III | 1.5 ± 2.1 | 2.0 ± 2.5 | 0.326 |
| ***b. Metal exposure measures*** |  |  |  |
| ***Overall exposure metrics*** |  |  | **< 0.001** |
| HrsW<sub>90</sub> (hours) | 0 ± 0 | 241 ± 200 | < 0.001 |
| E<sub>90</sub> (mg-days/m<sup>3</sup>) | 0.003 ± 0 | 2.3 ± 2.0 | < 0.001 |
| YrsW (y) | 0 ± 0 | 26.2 ± 10.9 | < 0.001 |
| ELT (mg-y/m<sup>3</sup>) | 0.001 ± 0.0003 | 1.1 ± 0.7 | < 0.001 |
| ***Overall blood metal levels*** |  |  | **< 0.001** |
| Ca (mg/L) | 57.8 ± 8.75 | 58.4 ± 9.16 | 0.766 |
| Cr (µg/L) | 7.91 ± 20.5 | 29.1 ± 148 | 0.102 |
| Cu (µg/L) | 749 ± 133 | 894 ± 124 | < 0.001 |
| Fe (mg/L) | 498 ± 76 | 556 ± 53 | 0.002 |
| K (mg/L) | 1765 ± 342 | 2085 ± 394 | 0.001 |
| Mg (mg/L) | 35.7 ± 5.17 | 38.4 ± 6.48 | 0.070 |
| Mn (µg/L) | 8.9 ± 2.5 | 11.0 ± 3.2 | 0.002 |
| Na (mg/L) | 2037 ± 370 | 2191 ± 389 | 0.134 |
| Pb (µg/L) | 8.5 ± 4.8 | 22.7 ± 18.5 | < 0.001 |
| Se (µg/L) | 149 ± 69.8 | 220 ± 105 | 0.001 |
| Sr (µg/L) | 13.9 ± 5.17 | 14.0 ± 6.26 | 0.928 |
| Zn (µg/L) | 5.62 ± 1.57 | 6.44 ± 1.47 | 0.026 |

Data represent the mean ± SD for each measure. Groups were compared using one-way analysis of variance (ANOVA). Metal exposure measures were compared using multivariate analysis of variance (MANOVA). Abbreviations: y = years; MoCA = Montreal Cognitive Assessment; UPDRS = Unified PD Rating Scale; HrsW<sub>90</sub> = Hours welding, 90 days; E<sub>90</sub> = Cumulative 90 day exposure to Mn; YrsW = Years welding, lifetime; ELT = Cumulative exposure to Mn, lifetime;

### Group comparison of medial temporal MRI and neuropsychological metrics

There were no group differences in morphological metrics including hippocampal volume and cortical thickness in entorhinal and parahippocampal cortices with age and education included as covariates (p’s> 0.117; Figure 3a-b). Exposed subjects (welders) displayed higher diffusivity (MD, AD, and RD) in all ROIs (p’s<0.040; Figure 4a-c). FA were significantly lower in entorhinal and parahippocampal cortices (p’s <0.033), but not in the hippocampus (p=0.501; Figure 4d).

**Figure 3.**
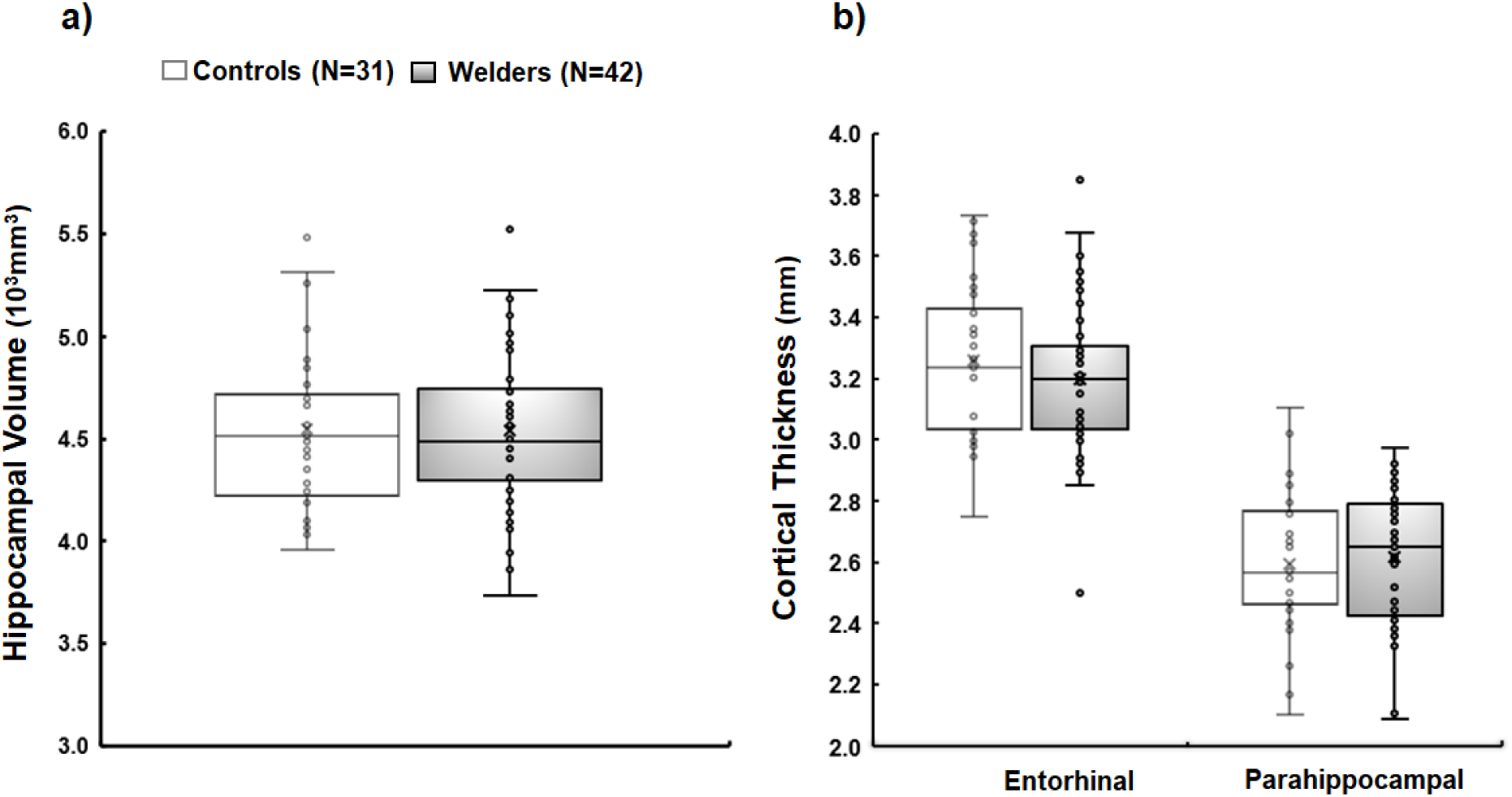
Morphologic MRI measures in exposed subjects (welders) and controls: a) hippocampal volume; b) cortical thickness in entorhinal and parahippocampal cortices.

**Figure 4.**
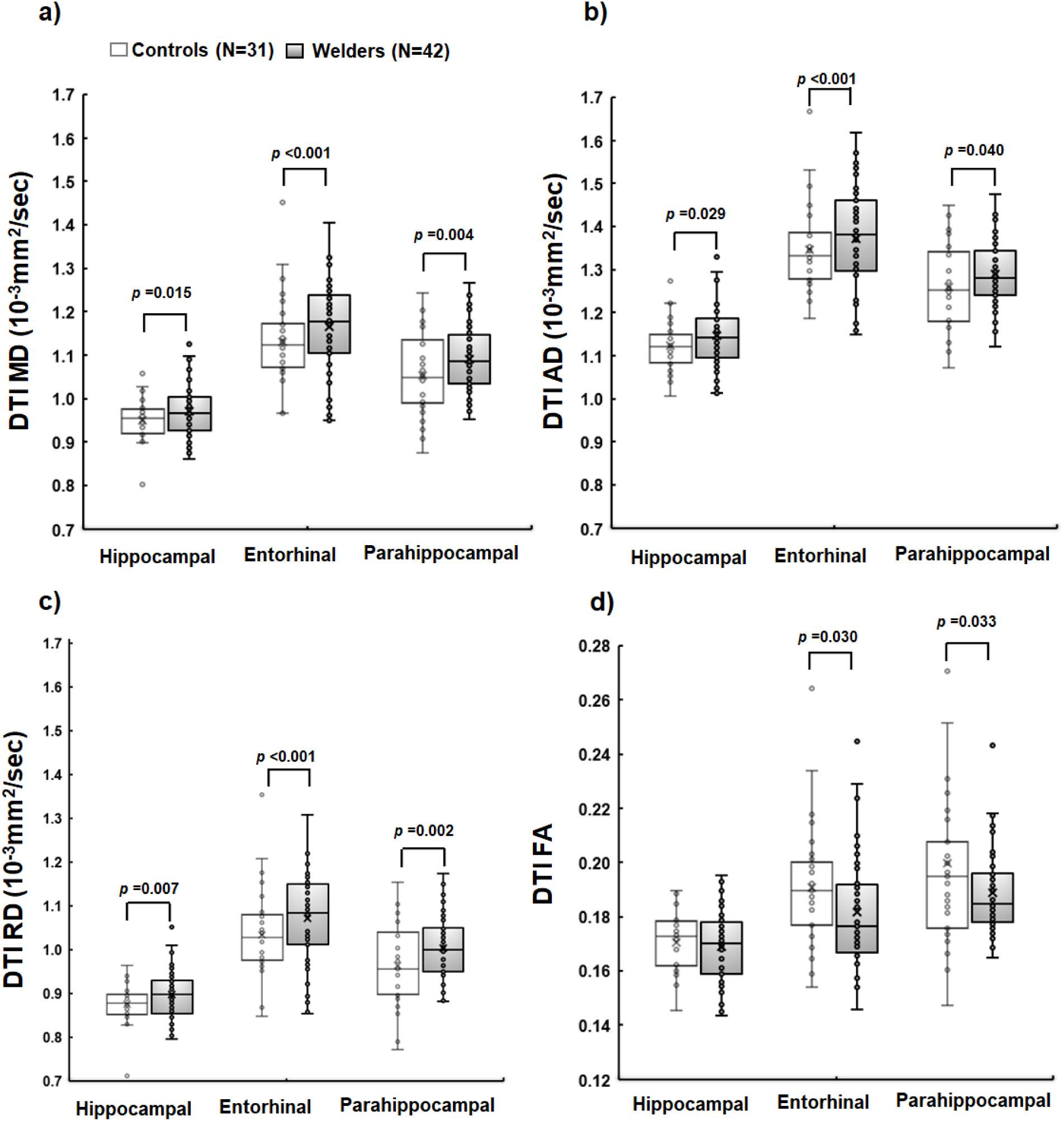
Diffusion tensor imaging (DTI) a) mean (MD); b) axial (AD), c) radial (RD) diffusivity, and d) fractional anisotropy (FA) of medial temporal structures for exposed subjects (welders) and controls.

Exposed subjects displayed overall lower cognitive performance than controls, but the group differences did not reach statistical significance (p=0.063; Table 2). <u>In domain-wise</u> <u>analyses</u>, exposed subjects performed worse in processing/psychomotor speed, executive function, and visuospatial processing than controls (p’s <0.046), but similarly on language (p=0.487) and attention/working memory (p=0.126) tests. Exposed subjects also performed poorer on learning/memory tasks, but the difference did not reach statistical significance (p=0.055). <u>For</u> <u>individual subtests</u>, exposed subjects performed worse on *Stroop-Color* and *Stroop-Word* processing/psychomotor speed and *Symbol-Digit Coding*, *Phonemic Fluency*, and *Trail Making-B* executive function tasks than controls. Exposed subjects also scored lower on *Complex Figure-Copy* visuospatial processing and *Delayed Story Recall* learning/memory tasks (p’s <0.049). There were no significant group differences in the remaining individual subtests (p’s >0.131).

**Table 2.** Neuropsychological test results for controls and exposed subjects.

| *Test* | *Controls*<br>*(n=31)* | *Exposed subjects*<br>*(n=42)* | *P-value* |
| --- | --- | --- | --- |
| **Overall Cognition .....** |  |  | **0.063** |
| **Processing/Psychomotor speed.....** |  |  | **0.046** |
| Stroop-Color<sup>b</sup> | -0.35 ± 1.16 | -0.80 ± 1.03 | **0.034** |
| Stroop-Word<sup>b</sup> | -0.03 ± 1.05 | -0.84 ± 1.19 | **0.004** |
| Symbol Search<sup>a</sup> | 0.88 ± 0.99 | 0.43 ± 0.67 | **0.011** |
| Trail Making-A<sup>b</sup> | 0.69 ± 0.79 | 0.50 ± 0.75 | 0.203 |
| **Executive function.....** |  |  | **<0.001** |
| Color-word<sup>b</sup> | 0.12 ± 0.58 | 0.06 ± 0.70 | 0.512 |
| Symbol-Digit Coding<sup>a</sup> | 0.46 ± 1.06 | -0.10 ± 0.78 | **0.010** |
| Phonemic Fluency<sup>b</sup> | 0.36 ± 0.87 | -0.31 ± 0.73 | **<0.001** |
| Trail Making-B<sup>b</sup> | 0.51 ± 0.83 | -0.35 ± 1.41 | **0.004** |
| **Language .....** |  |  | **0.487** |
| Picture Naming<sup>a</sup> | 0.24 ± 1.13 | 0.59 ± 0.24 | 0.468 |
| Semantic Fluency<sup>a</sup> | 0.04 ± 1.06 | -0.23 ± 0.99 | 0.365 |
| **Visuospatial Processing.....** |  |  | **0.045** |
| Judgement of Line Orientation<sup>a</sup> | 0.69 ± 0.94 | 0.55 ± 0.82 | 0.858 |
| Complex Figure Copy<sup>a</sup> | -0.22 ± 0.86 | -1.09 ± 1.35 | **0.013** |
| **Learning/Memory .....** |  |  | **0.055** |
| List Learning<sup>a</sup> | -0.64 ± 0.93 | -0.76 ± 1.00 | 0.493 |
| Immediate Story Recall<sup>a</sup> | 0.06 ± 0.93 | -0.42 ± 1.04 | 0.151 |
| Delayed List Recall<sup>a</sup> | -0.73 ± 0.98 | -0.52 ± 1.01 | 0.192 |
| Delayed Story Recall<sup>a</sup> | 0.13 ± 0.90 | -0.46 ± 1.04 | **0.049** |
| Delayed Figure Recall<sup>a</sup> | -0.13 ± 1.10 | 0.01 ± 1.16 | 0.261 |
| **Attention/working memory .....** |  |  | **0.126** |
| Letter Number Sequencing<sup>a</sup> | 0.30 ± 1.00 | 0.03 ± 0.94 | 0.906 |
| Digit Span<sup>a</sup> | 0.81 ± 1.11 | 0.09 ± 0.83 | 0.131 |
| Spatial Span<sup>a</sup> | 0.34 ± 0.65 | -0.10 ± 0.93 | 0.176 |

Data represent the mean ± SD. Test scores were converted to z-scores based on age- (denoted as <sup>a</sup>) or age- and education- (denoted as <sup>b</sup>) adjusted norm (t- or scaled) scores. Higher scores indicate better performance. Multivariate analysis of covariance (MANCOVA) was conducted with Group as the independent variable and individual subtests of each cognitive domain as the dependent variables. Bold numbers indicate significant result at $p < 0.05$ . Education level was additionally adjusted for the cognitive domains language, visuospatial processing, learning/memory, and attention/working memory.

### The associations among whole blood metal levels in exposed subjects

In exposed subjects, greater blood Cu levels correlated with greater blood K, Pb, and Se (p’s <0.002, Supplementary Table S1). Greater blood Fe additionally correlated with greater blood K, Mn, and Zn (p’s <0.002). Greater blood K correlated with greater Pb, Se, and Zn (p’s<0.001). Greater blood mg additionally correlated with greater blood Mn, Pb, Se, and Zn (p’s <0.008). Greater blood Pb additionally correlated with blood Se (p=0.005). Greater blood Se additionally correlated with blood Zn (p<0.001). These correlations remained significant after multiple comparison correction (Supplementary Table S1).

### The associations of metal exposure with MRI and neuropsychological metrics

#### Mixed-metal exposure history as the exposure of interest

##### Diffusion MRI

Pearson partial correlation analyses showed that lower parahippocampal FA correlated with greater YrsW (R=-0.386, p=0.013). Higher hippocampal RD values were associated with greater ELT values (R=0.317, p=0.046). DTI values in other ROIs did not correlate with any other mixed-metal exposure history (p’s >0.062; Supplementary Table S2). Subsequent multiple linear regression analysis confirmed that greater YrsW was a significant predictor of lower parahippocampal FA (ß=-0.001, t=-2.61, p=0.013; Table 3 and Figure 5a). Higher ELT was a significant predictor of higher hippocampal RD (ß=0.021, t=2.06, p=0.046).

**Figure 5.**
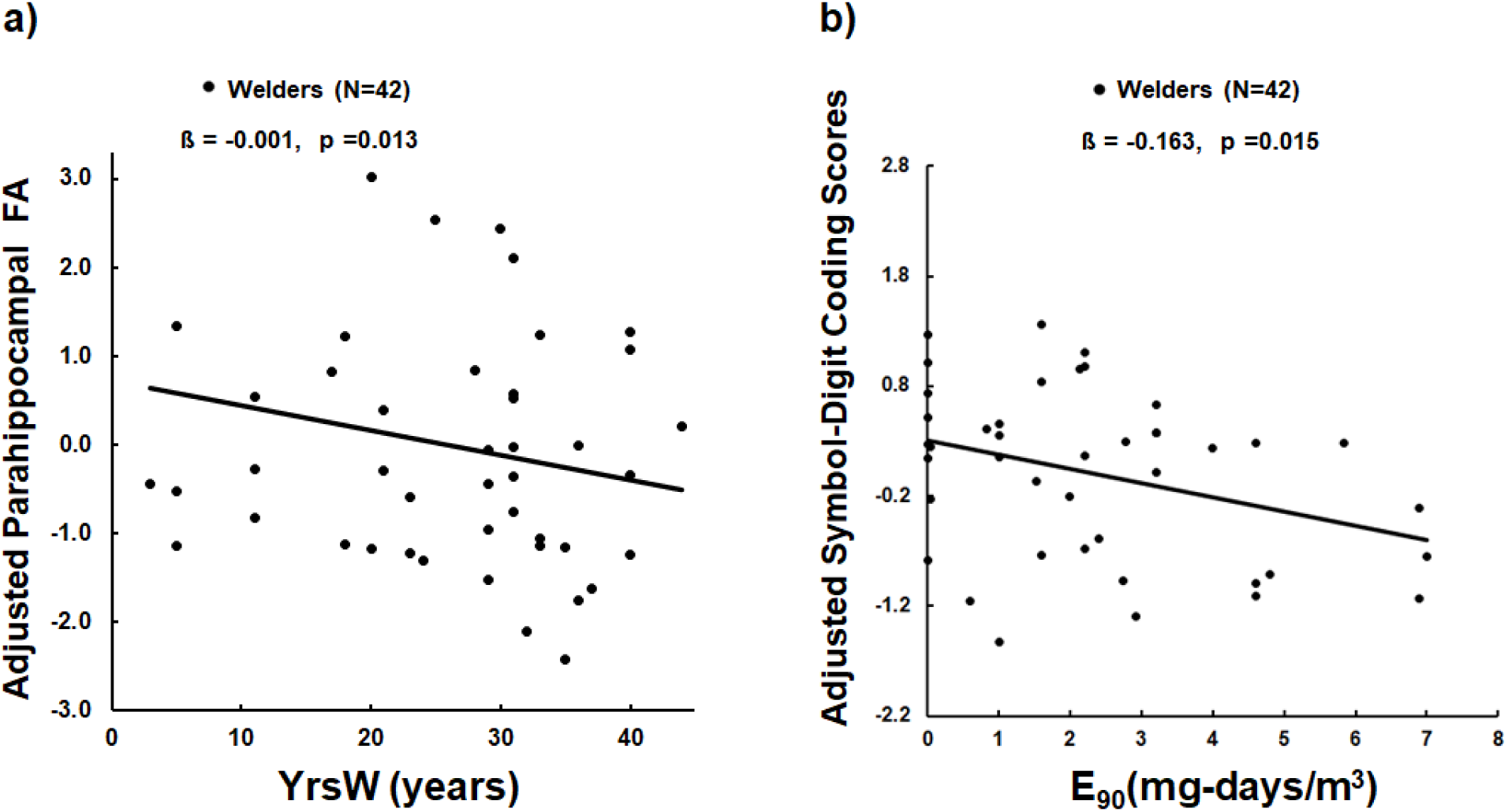
Scatter plots show a) adjusted parahippocampal FA values (y-axis) vs. YrsW (x-axis) in exposed subjects (welders); b) adjusted *Symbol-Digit Coding* scores (y-axis) vs. E_90_ (x-axis). Adjusted values indicate values after controlling for age and education level.

**Table 3.**
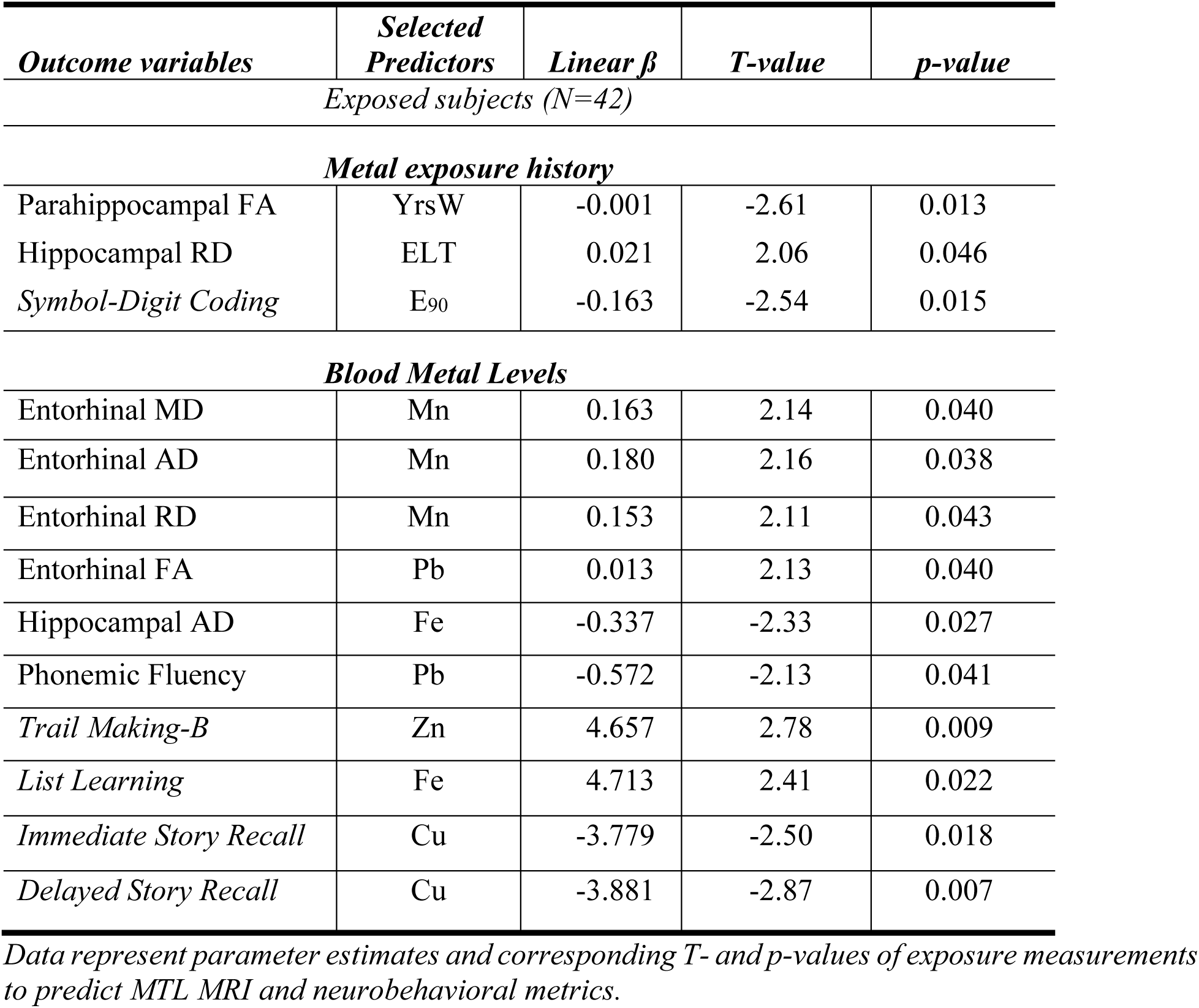
Multiple linear regression analyses evaluating the influence of exposure measures on medial temporal MRI and neurobehavioral metrics for exposed subjects.

##### Neuropsychological tests

Pearson correlation analyses revealed that lower *Symbol-Digit Coding* scores correlated with higher E_90_ (R=-0.381, p=0.015; Supplementary Table S3). The remaining neuropsychological scores did not correlate with any other mixed-metal exposure history (p’s >0.062). Multiple linear regression analysis (Table 3) revealed that higher E_90_ was a significant predictor of lower *Symbol-Digit Coding* scores (ß=-0.163, t=-2.54, p=0.015; Figure 5b).

#### Whole blood metal levels as the exposure of interest

##### Diffusion MRI

Pearson partial correlation analyses showed that higher entorhinal diffusivities correlated with higher Mn (R’s >0.341, p’s <0.049, Supplementary Table S2). Higher entorhinal FA values correlated with higher Pb (R=0.348, p=0.040). None of the DTI metrics in other ROIs correlated with any blood metal levels (p’s>0.067). Subsequent multiple linear regression analysis (Table 3) revealed that higher Mn values were significant predictors of higher entorhinal diffusivities (ß’s>0.153, t’s>2.11, p’s <0.043). Higher Pb significantly predicted higher entorhinal FA values (ß=0.013, t=2.13, p=0.040), whereas higher Fe level predicted lower hippocampal AD (ß=-0.337, t=-2.33, p=0.027).

##### Neuropsychological tests

Pearson correlation analyses revealed that lower *Phonemic Fluency* scores were associated with higher blood Pb (R=-0.358, p=0.041, Supplementary Table S3). In contrast, lower *Trail Making-B* and *List Learning* scores were associated with lower blood Zn (R=0.449, p=0.009) and Fe (R=0.397, p=0.022), respectively. In addition, poorer *Immediate* and *Delayed Story Recall* scores correlated with higher blood Cu levels (R=-0.410, p=0.018 and R=-0.458, p=0.007, respectively). The *Delayed Story Recall*-blood Cu association remained significant after *FWER* correction. The remaining neuropsychological test scores did not correlate with any other blood metal levels (p’s >0.080). Multiple linear regression analyses (Table 3) revealed that higher blood Pb levels were a significant predictor of poorer *Phonemic Fluency* (ß=-0.572, t=-2.13, p=0.041). Lower blood Zn and Fe were significant predictors of lower *Trail Making-B* and *List Learning* scores (ß=4.657, t=2.78, p=0.009 for Zn; ß=4.713, t=2.41, p=0.022 for Fe). Higher blood Cu was a significant predictor of poorer *Immediate* and *Delayed Story Recall* (ß=-3.779, t=-2.50, p=0.018 and ß=-3.881, t=-2.87, p=0.007, respectively; Figure 6d).

**Figure 6.**
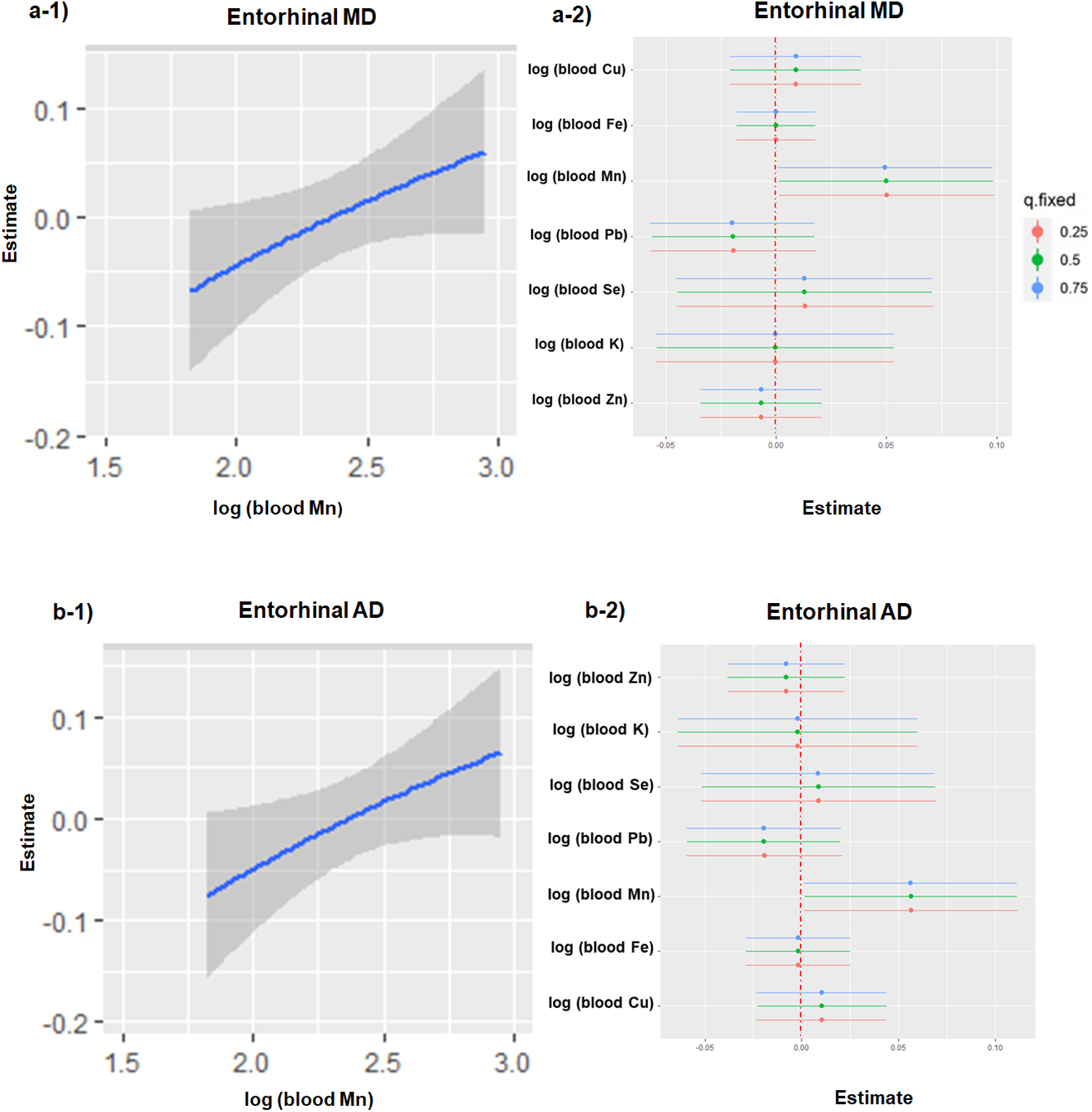
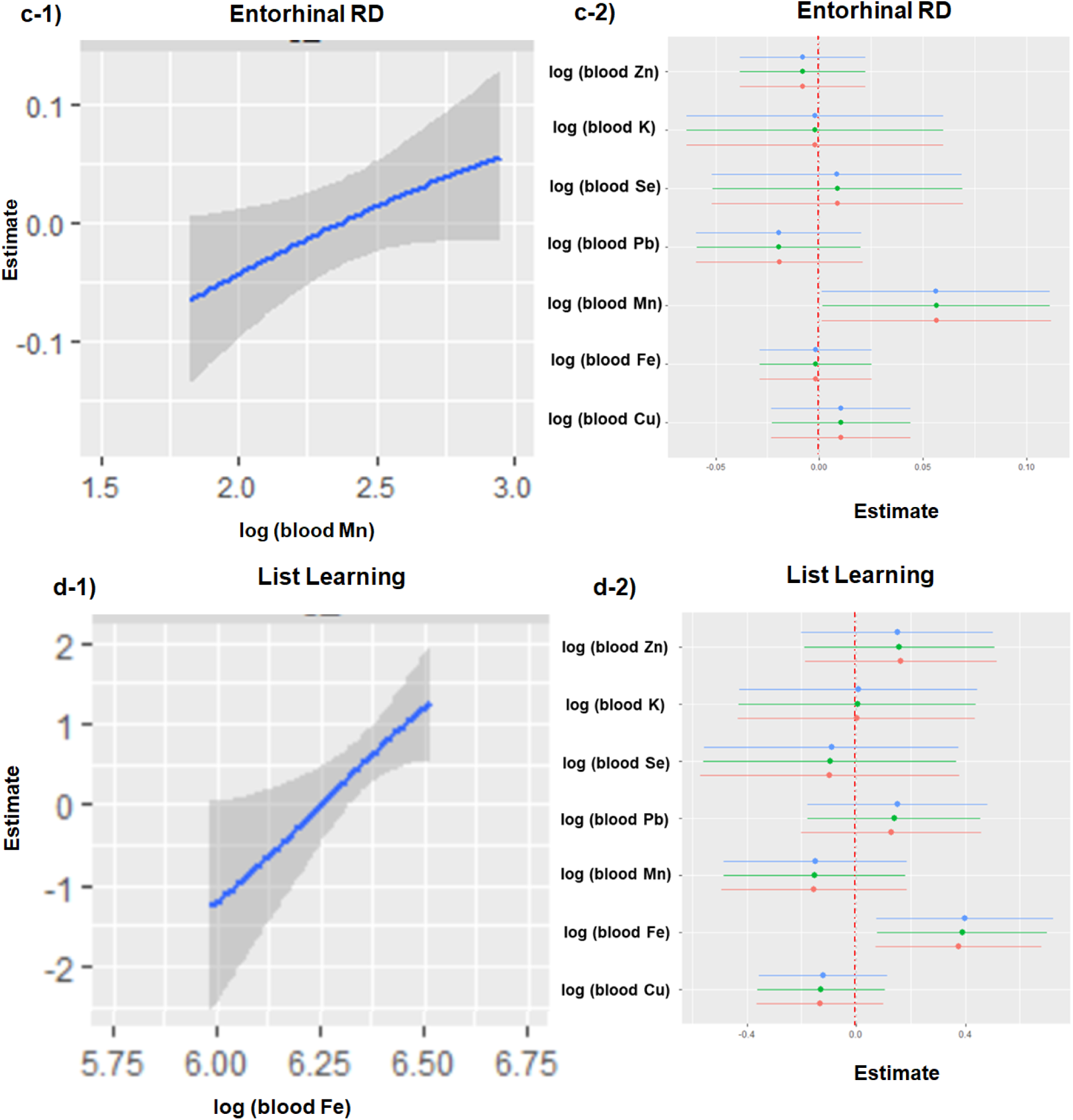
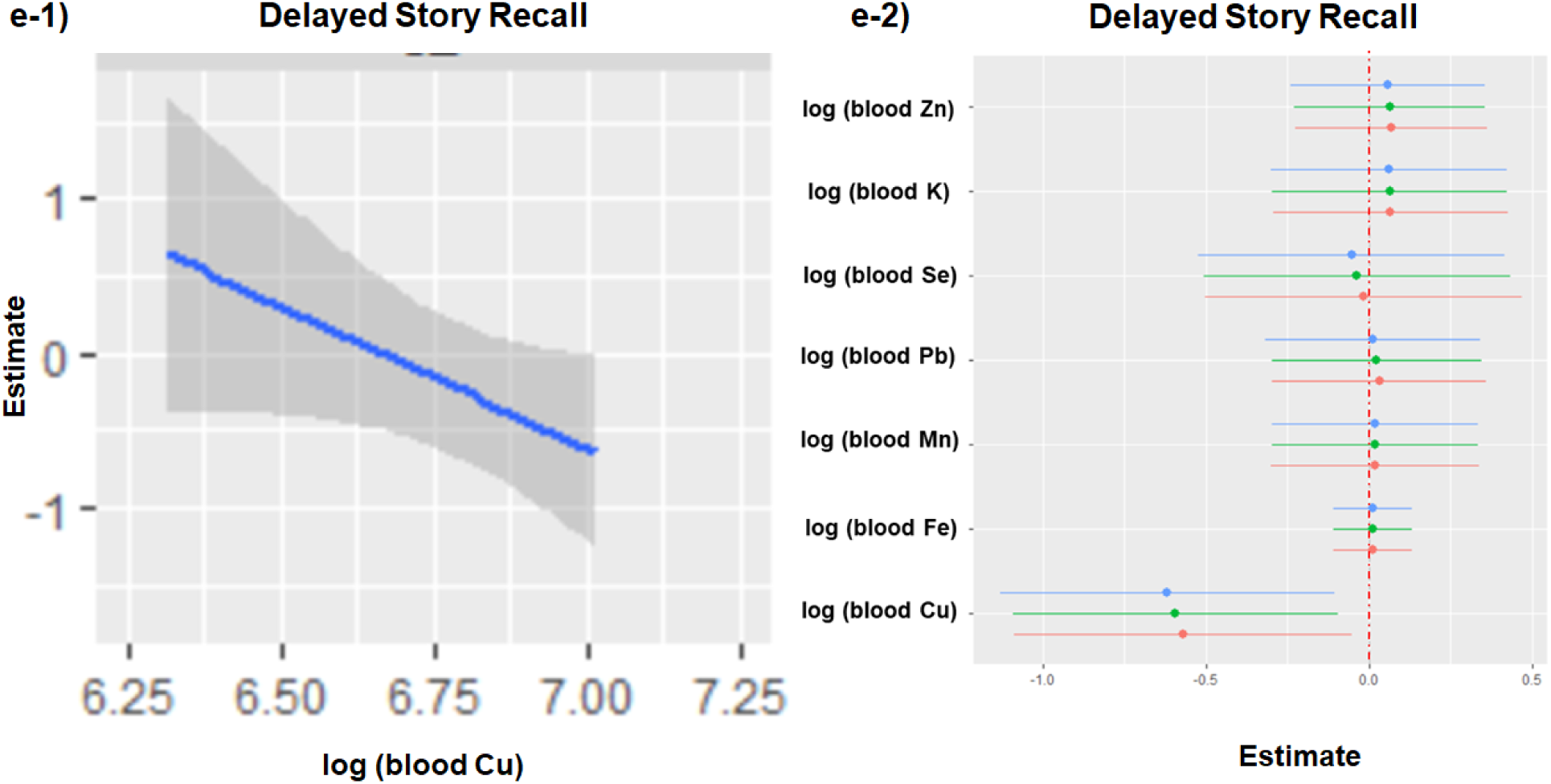
BKMR (Bayesian kernel machine regression) univariate dose-response associations of metal mixture components with medial temporal DTI and neuropsychological scores with 95% confidence interval band in exposed subjects; a-1) estimate of metal exposure dose-response function (y-axis) vs. log-transformed blood Mn (x-axis) for entorhinal MD; a-2) log-transformed whole blood metals at different quantiles (y-axis) vs. estimate of metal exposure dose-response function (x-axis) for entorhinal MD; b-1) estimate of metal exposure dose-response function (y-axis) vs. log-transformed blood Mn (x-axis) for entorhinal AD; b-2) log-transformed whole blood metals at different quantiles (y-axis) vs. estimate of metal exposure dose-response function (x-axis) for entorhinal AD; c-1) estimate of metal exposure dose-response function (y-axis) vs. log-transformed blood Mn (x-axis) for entorhinal RD; c-2) log-transformed whole blood metals at different quantiles (y-axis) vs. estimate of metal exposure dose-response function (x-axis) for entorhinal RD; d-1) estimate of dose-response function (y-axis) vs. log-transformed blood Fe (x-axis) for *List Learning* scores; d-2) log-transformed whole blood metals at different quantiles (y-axis) vs. estimate of metal exposure dose-response function (x-axis) for *List Learning* scores; e-1) estimate of metal exposure dose-response function (y-axis) vs. log-transformed blood Cu (x-axis) for *Delayed Story Recall* scores; e-2) log-transformed whole blood metals at different quantiles (y-axis) vs. estimate of metal exposure dose-response function (x-axis) for *Delayed Story Recall* scores. Age and education level were used as covariates. Other blood metal levels than the metal of interest in the analysis were fixed at the median on the y-axis.

##### Overall metal mixture and interaction effects using BKMR analyses

When examining effects of single exposure predictors on brain DTI and neuropsychological metrics, higher blood Mn at different quartiles (e.g., 25^th^, 50^th^, and 75^th^) was a significant predictor of higher entorhinal AD, MD, and RD values (p’s <0.05; Figure 6a-c). Higher blood Fe levels at different quartiles (e.g., 25^th^, 50^th^, and 75^th^) were a significant predictor of better *List Learning* (p<0.05; Figure 6d), whereas higher blood Cu levels at different quartiles (e.g., 25^th^, 50^th^, and 75^th^) were a significant predictor of poorer *Delayed Story Recall* (p<0.05; Figure 6e). None of the blood metal levels were significant predictors of hippocampal DTI metrics.

When examining overall effects of metal mixture by comparing estimated dose-response function of all predictors at a particular percentile (e.g., from 25^th^ to 75th) compared to that of at their 50^th^ percentile, there also were no significant overall metal mixture effects at different quartiles (e.g., from 25^th^ to 75th) on MTL DTI or neuropsychological metrics (p’s >0.05; Supplementary Figure S1). When examining pairwise interaction effects among multiple metals by testing a single exposure predictor-response function while a second exposure predictor fixed at different quantiles (e.g., 25^th^, 50^th^, and 75^th^) and remaining exposure predictors fixed at median values, none of pairwise interactions among multiple metals were significant (p’s>0.05; Supplementary Figure S2).

### The associations between MTL DTI and neuropsychological test scores

#### Pearson partial correlation analyses (Supplementary Table S3)

Higher entorhinal diffusivities correlated with poorer Delayed List Recall (R’s<-0.415, p’s<0.010). These associations remained significant after FWER correction. Higher entorhinal MD additionally correlated with poorer List Recall (R=-0.320, p=0.044). Higher parahippocampal diffusivities correlated with poorer Immediate Story Recall (R’s<-0.432, p’s<0.005) and these associations remained significant after FWER correction. Lower parahippocampal FA correlated with lower Symbol Search and Trail Making-A scores (R’s>0.344, p’s<0.030). Higher hippocampal AD correlated with poorer List Learning (R=-0.334, p’s<0.025). Other DTI metrics did not correlate with any neurobehavioral scores (p’s>0.062).

#### Multiple linear regression analyses

Higher entorhinal diffusivities were significant predictors of *Delayed List Recall* scores (ß’s<-3.594, t’s<-2.74, p’s <0.010; Table 4 and Figure 7a). Higher parahippocampal diffusivities and higher entorhinal MD were significant predictors of poorer *Immediate Story Recall* scores (ß’s<-3.426, t’s<-2.08, p’s <0.044; Figure 7b). Lower parahippocampal FA values were a significant predictor of lower *Symbol Search* and *Trail Making-A* scores (ß=16.058, t=2.39, p=0.022 and ß=16.908, t=2.26, p=0.030, respectively; Figure 7c-d). Higher hippocampal AD values were a significant predictor of lower *List Learning* scores (ß=-4.728, t=-2.18, p=0.035; Figure 7e).

**Figure 7.**
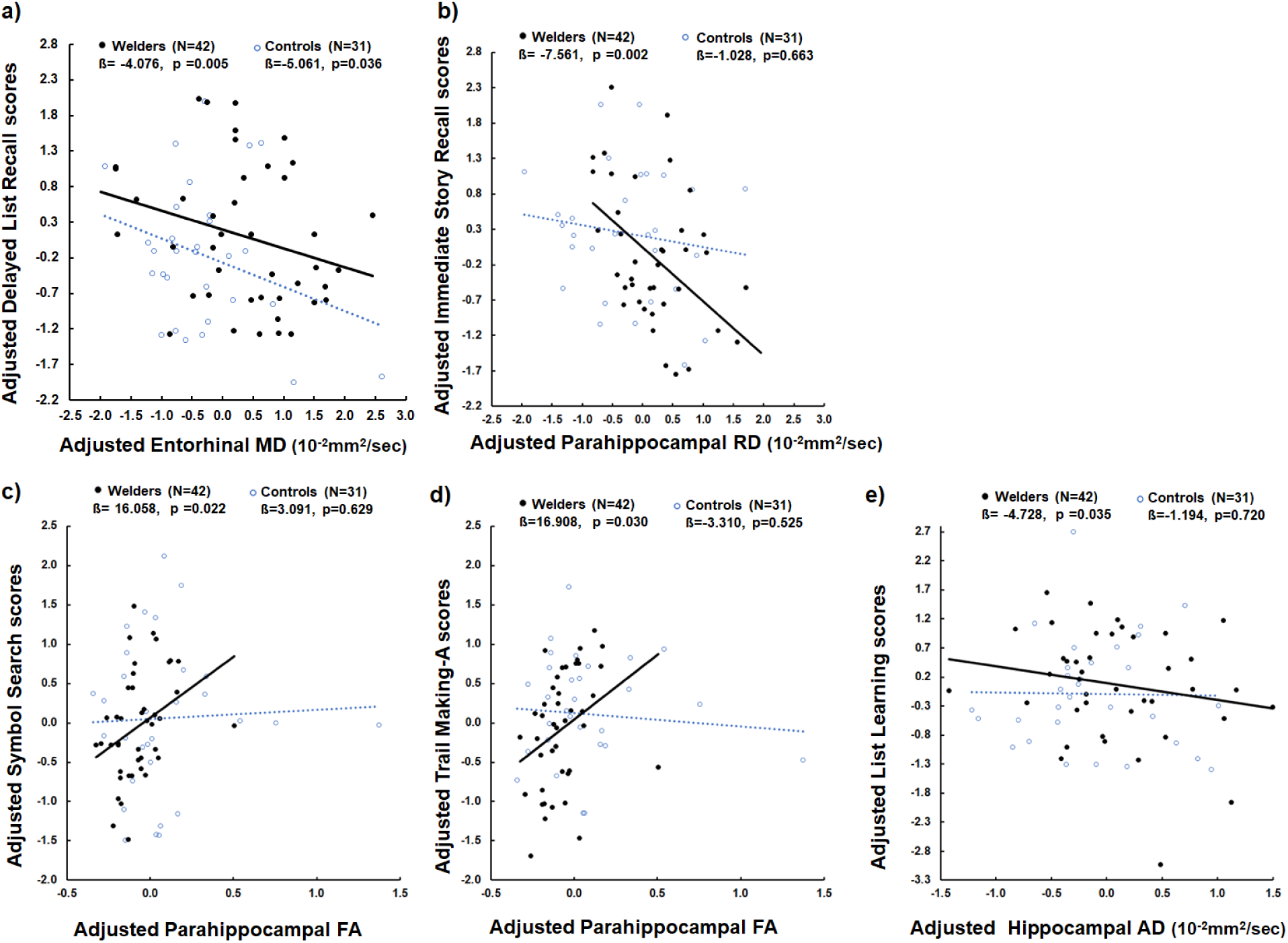
Scatter plots show a) adjusted *Delayed List Recall* scores (y-axis) vs. adjusted entorhinal MD values (x-axis); b) adjusted *Immediate Story Recall* scores (y-axis) vs. adjusted parahippocampal RD values (x-axis); c) *Symbol Search* scores (y-axis) vs. adjusted parahippocampal FA values (x-axis); d) adjusted *Trail Making-A* scores (y-axis) vs. adjusted parahippocampal FA values (x-axis); e) adjusted *List Learning* scores (y-axis) vs. adjusted hippocampal AD values (x-axis) in both exposed subjects (welders) and controls.

**Table 4.**
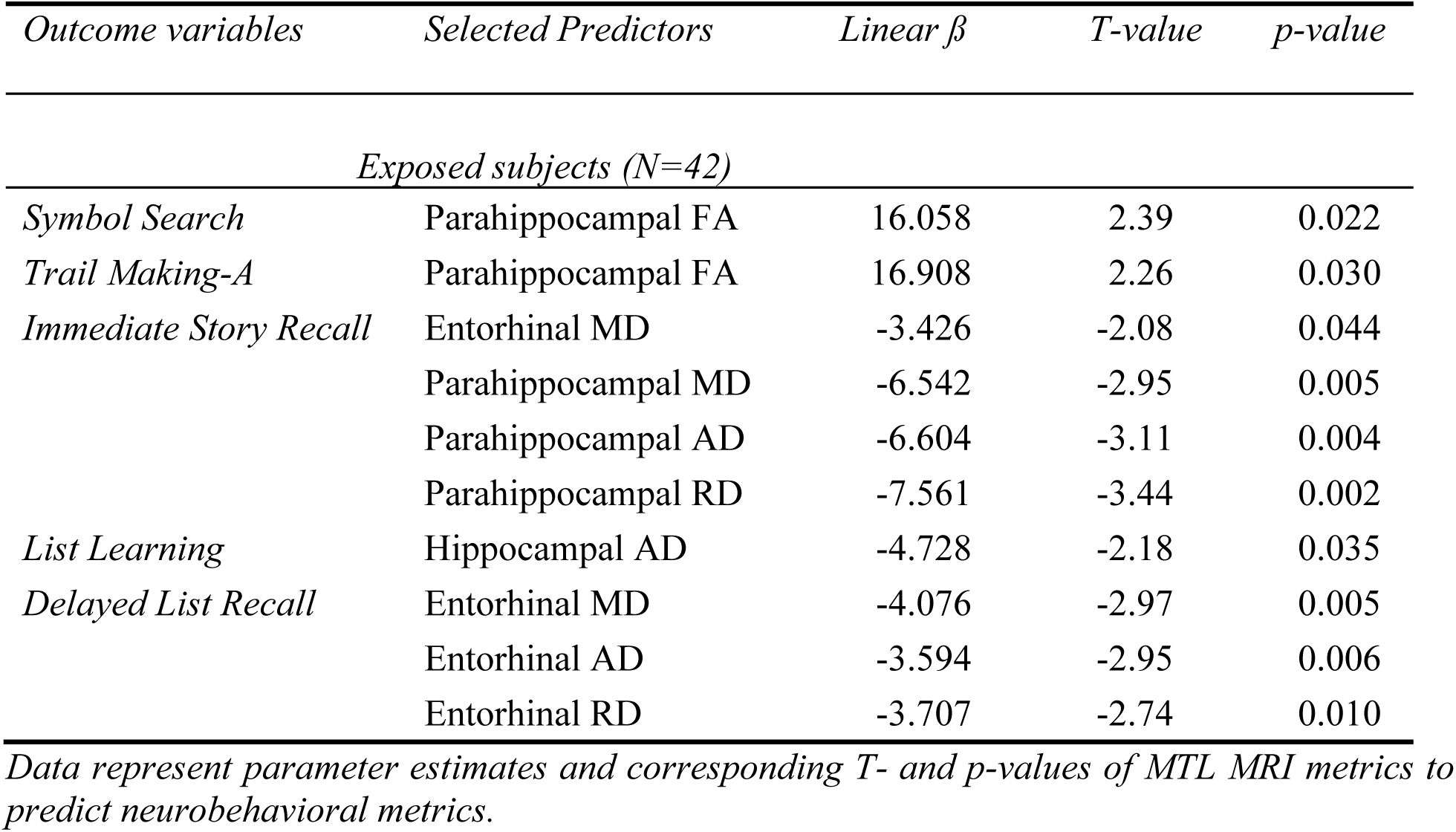
Multiple linear regression analyses to determine medial temporal structural MRI predicting neurobehavioral metrics for exposed subjects.

| *Outcome variables* | *Selected Predictors* | *Linear $\beta$* | *T-value* | *p-value* |
| --- | --- | --- | --- | --- |
| *Exposed subjects (N=42)* |  |  |  |  |
| *Symbol Search* | Parahippocampal FA | 16.058 | 2.39 | 0.022 |
| *Trail Making-A* | Parahippocampal FA | 16.908 | 2.26 | 0.030 |
| *Immediate Story Recall* | Entorhinal MD | -3.426 | -2.08 | 0.044 |
|  | Parahippocampal MD | -6.542 | -2.95 | 0.005 |
|  | Parahippocampal AD | -6.604 | -3.11 | 0.004 |
|  | Parahippocampal RD | -7.561 | -3.44 | 0.002 |
| *List Learning* | Hippocampal AD | -4.728 | -2.18 | 0.035 |
| *Delayed List Recall* | Entorhinal MD | -4.076 | -2.97 | 0.005 |
|  | Entorhinal AD | -3.594 | -2.95 | 0.006 |
|  | Entorhinal RD | -3.707 | -2.74 | 0.010 |

*Data represent parameter estimates and corresponding T- and p-values of MTL MRI metrics to predict neurobehavioral metrics.*

### Causal mediation analyses: from exposure to neurobehavior via MTL MRI feature

#### Mixed-metal exposure history as the exposure of interest

There were significant indirect effects of higher YrsW on lower *Symbol Search* and *Trail Making-A* scores that were mediated by lower parahippocampal FA values (ß=-0.025, z=-2.29, p=0.022 for *Symbol Search* and ß=-0.028, z=-2.27, p=0.023 for *Trail Making-A*) but no significant direct or total effects (p’s>0.055). There were significant total and direct effects of higher E_90_ values on lower *Symbol-Digit Coding* scores (ß’s<-0.153, z’s<-2.44, p’s<0.014) but no significant indirect effects (p’s>0.647). There were no significant direct or indirect effects of other mixed-metal exposure history on neuropsychological scores (p’s>0.065).

#### Whole blood metal levels as the exposure of interest

There were significant total (ß=6.401, z=3.85, p<0.001) and indirect effects of higher blood Fe on higher *List Learning* scores via lower hippocampal AD (ß=7.567, z=2.11, p=0.035) without significant direct effects (p=0.726). Blood Mn levels had significant total effects (ß’s<-1.230, z’s <2.24, p’s <0.025) on *Delayed List Recall* scores without significant direct or indirect effects via MTL DTI metrics (p’s>0.109). Blood Cu levels had significant total and direct effects (ß’s<-3.1633, z’s<-1.97, p’s<0.049) on lower *Symbol-Digit Coding* and *Delayed Story Recall* scores without significant indirect effects via MTL DTI metrics (p’s>0.366). Blood Pb levels had significant total and direct effects (ß’s<-0.614, z’s<-2.23, p’s<0.026) on lower *Phonemic Fluency* scores without significant indirect effects via MTL DTI metrics (p’s>0.366).

## Discussion

This is the first in-depth examination of the relationship between mixed metal exposure and brain structural features in several MTL regions using multimodal MRI along with neuropsychological features. Consistent with our hypothesis H1, we observed a reliable pattern differences of MRI diffusion features (higher AD, RD, and MD and lower FA) across MTL areas that is consistent with findings in Alzheimer’s disease at-risk populations. Along with the absence of morphometric differences (markers of significant macroscopic neuronal loss), these findings suggest that these MTL DTI metrics may serve as sensitive biomarkers for gauging early pathological processes that may reflect microstructural integrity. Consistent with our hypothesis H2, subjects with mixed metal exposure had lower performance in the select processing/psychomotor speed and executive function domains, as well as the *Delayed Story Recall* task, despite having no detectable deficits in the broader learning/memory domain.

This is also the first comprehensive and systematic analyses of metal exposure dose-response (brain-related health outcomes: MRI and neuropsychological metrics) analyses including correlations, multiple linear regression, and BKMR analyses. First, our data showed that long-term mixed-metal exposure history (YrsW and ELT) predicted altered MTL DTI metrics (lower parahippocampal FA and higher hippocampal RD). Consistent with hypothesis H3, we also observed robust associations of higher blood Mn and Cu levels with higher entorhinal diffusivity, as well as with lower *Delayed Story Recall* performance. These features are often seen in populations with Alzheimer’s disease and related disorders. There were, however, no significant direct (?) or interaction effects of metal mixtures as measured in blood on MTL MRI or cognitive performance. MTL DTI metrics (microstructural differences) that would mediate, at least partially, the effects of mixed metal exposure (measure as blood levels) on cognitive performance.

Together, these results support that mixed metal exposure is associated with MTL microstructural and neuropsychological features resembling Alzheimer’s disease at-risk populations. Given the ubiquitous nature of environmental metals and growing public health concerns of its link to Alzheimer’s disease, follow-up studies are warranted and may help to develop and implement effective prevention strategies against Alzheimer’s disease and other age-related neurogenerative disorders.

### MTL features resemble Alzheimer’s disease at-risk population in exposed subjects

The hippocampus and entorhinal/parahippocampal cortices are key structures for learning/memory tasks. The parahippocampal gyrus receives converging inputs from widespread associative areas including frontal, temporal, parietal, occipital, and cingulate cortices (Aminoff et al., 2013; Suzuki, 2009). It then projects to the entorhinal cortex that, in turn, has reciprocal connections with the hippocampus. Thus, the entorhinal cortex serves as a major information relay station to the hippocampus and from the hippocampus back to the cortical association areas via the parahippocampal gyrus (Raslau et al., 2015; Wixted and Squire, 2011). These pathways parallel the fimbria-fornix system of the hippocampal projections to diencephalic and other limbic system structures subserving particularly memory consolidation (Eichenbaum and Lipton, 2008). The entorhinal outputs then are projected back to various cortical regions through the parahippocampal cortex.

Consistent with its role in learning and memory, MTL areas appear to be among the first regions affected by Alzheimer’s disease-related pathological changes (Braak and Braak, 1991; Devanand et al., 2007; Echavarri et al., 2011; Scahill et al., 2002). Decreased hippocampal and entorhinal volume and/or thickness have been reported as predictors for conversion to Alzheimer’s disease (Apostolova et al., 2006; De Santi et al., 2001). Recent studies suggested other imaging metrics such as shape, texture, functional networks, or water molecule diffusion in neuronal tissues may be even more sensitive in capturing early Alzheimer’s disease-related alterations (Douaud et al., 2013; Kantarci et al., 2005; Leandrou et al., 2020; Leandrou et al., 2018; Nir et al., 2013; Weston et al., 2020). Our finding of altered MTL DTI features in exposed subjects is consistent with the idea that MTL is vulnerable to environmental neurotoxic metal exposures. More importantly, the fact that our exposed subjects were clinically asymptomatic and revealed no apparent MTL volume reduction or cortical thinning suggest that determining single or mixed metal exposure early may decrease the progression to more frank symptoms and eventually Alzheimer’s disease or ADRD.

### Neuropsychological features resembling Alzheimer’s disease at-risk population in exposed subjects

Subjects with chronic metal exposure are known to perform worse on several neuropsychological tests (Akila et al., 1999; Chang et al., 2009; Meyer-Baron et al., 2013). Alzheimer’s disease and related disorder-related neuropsychological deficits comprise various cognitive functions including executive, attentional, and language functions. Difficulties with learning/memory tasks, however, have the most robust association with early Alzheimer’s disease and related disorder (Backman et al., 2001; Tromp et al., 2015). Our finding that exposed subjects have lower *Delayed Story Recall* performance, rather than other types of episodic memory tasks (e.g., word list or visuospatial figure recall) suggests that Alzheimer’s disease and related disorder -related early episodic memory decline may occur in a task-specific way.

Episodic memory is a crucial function of the hippocampus (Tulving and Markowitsch, 1998). MTL arears, such as parahippocampal and entorhinal cortices, are known provide major inputs to the hippocampus (Eichenbaum and Lipton, 2008). Previous studies reported that lower entorhinal cortex volume, but not hippocampal volume, was associated with lower *Delayed List* and *Story Recall* scores in Alzheimer’s patients (Di Paola et al., 2007; Eichenbaum and Lipton, 2008). Furthermore, MTL DTI metrics have been associated with memory scores in both healthy elderly and Alzheimer’s patients (Carlesimo et al., 2010; Mayo et al., 2018). Together, these prior studies suggest MTL involvement in episodic memory and early Alzheimer’s disease process (Di Paola et al., 2007). Our findings of robust associations of higher entorhinal and parahippocampal diffusivities with lower (worse) learning/memory performance are consistent with this notion.

In the current study, our exposed subjects also displayed lower processing/psychomotor speed performance. Previous studies also observed significant associations between parahippocampal gyrus thickness and processing speed scores, particularly when working with visuospatial materials (Kraft et al., 2020; Takahashi et al., 2002). Speedy information processing may be crucial for various cognitive functions requiring attention, encoding and retrieval of to-be-remembered information, reasoning, decision making, and visuospatial perceptions (Salthouse, 1996). Given previously observed associations between slowed psychomotor speed and risk for dementia development (Andriuta et al., 2019; Bailon et al., 2010; Kuate-Tegueu et al., 2017), our results suggest neural correlates responsible for Alzheimer’s disease-related slowed psychomotor speed. Collectively, our data are consistent with that mixed-metal exposure is associated with cognitive deficits/patterns reported in Alzheimer’s disease at-risk populations (Backman et al., 2001; Douaud et al., 2013).

### From mixed metal exposure to MTL MRI and neuropsychological performance

Recent studies emphasize the importance of considering individual and collective roles of metal mixtures in neurotoxic processes (Saxena et al., 2022; Wu et al., 2023). In the present study, we utilized questionnaire-based instruments to assess mixed metal exposure history and whole blood metal measurements to estimate metal exposure dose and multiple statistical methods to examine complex dynamics of metal mixtures with MTL MRI and neuropsychological performance.

First, we found that long-term metal exposure history (e.g., ELT and YrsW) was significant predictors of hippocampal and parahippocampal DTI metrics, validating that MTL microstructural features are related to long-term metal exposure. Subsequent mediation analyses suggested that YrsW indirectly affected lower processing/psychomotor speed performance (lower *Symbol Search* and *Trail Making-A* scores) via lower parahippocampal FA values.

Second, we noted that short-term metal exposure history that takes exposure intensity into account (e.g., E_90_) predicted lower *Symbol-Digit Coding* scores, an executive function task where subjects matched symbols to designated numbers under time pressure. This task can be demanding for elderly adults because it requires executive function skills in addition to appropriate psychomotor speed. This result suggests that even short-term metal exposure history when coupled with higher intensity may interfere with proper performance on executive function tasks.

Third, higher whole blood metal levels (e.g., blood Mn and Cu), surrogate markers for short-term metal exposure, demonstrated robust associations with higher entorhinal diffusivity and lower episodic memory performance (lower *Delayed Story Recall* scores) that persisted throughout multiple statistical methods of correlation, multiple linear regression, and BKMR analyses. The finding of whole blood Mn-entorhinal diffusivity association is novel. Note that a significant association between whole blood Mn and DTI metrics (e. g., lower FA) was previously reported in metal-exposed subjects (e.g., welders) (Kim et al., 2011). In that study, however, regions of interest were limited to white matter structures (e.g., corpus callosum and frontal white matter areas) and did not include MTL structures. In addition, mediation analyses suggested a significant link between higher blood Mn and lower episodic memory performance (lower *Delayed List Recall* scores) but the exact characteristic of the link (direct or indirect via MTL microstructural features) was inconclusive. This may be due partly to the fact that a whole blood metal marker may not sensitively reflect long-term exposure effects.

The significant association between higher whole blood Cu and lower episodic memory (lower *Delayed Story Recall* scores) in subjects with multiple metal exposure is also novel. Chronic Cu exposure has been associated with increased brain Cu accumulation in the basal ganglia and hippocampus(Pal et al., 2013) and increased risk for Alzheimer’s disease (Patel and Aschner, 2021). Cu in brain is known to have a high affinity for β-amyloid plaques that may enhance inflammatory responses, reactive oxygen species (ROS) generation, and fibril formation leading to increased β-amyloid aggregates, a characteristic feature of Alzheimer’s disease and related disorders (Bagheri et al., 2017). Future environmental health studies need to include Cu as an important risk factor for neurodegenerative disorders.

### Lessons learned from our comprehensive analyses

As the first comprehensive and systematic analyses of mixed metal exposure dose-response relationship using MRI and neuropsychological metrics as brain-related health outcomes, we also made several intriguing observations as discussed below that are worthwhile for future investigations. First, higher whole blood Fe levels in exposed subjects predicted a better *List Learning* score, yet there was no group difference in this task implying that Fe-related beneficial effects on cognition were overall not significant. Subsequent mediation analyses suggested that higher whole blood Fe is linked to higher *List Learning* scores via lower hippocampal AD values. These results suggest that higher peripheral Fe may be a protective factor against microstructural changes in the brain that lead to beneficial behavioral effects. This finding appears contradictory to previously reported neurotoxic effects of Fe exposure (Salvador et al., 2010; Yarjanli et al., 2017), yet it is consistent with reports of a complex association between peripheral Fe levels and cognitive tasks (Gong et al., 2021; Hosking et al., 2018; Li et al., 2019b). Fe is most abundant trace metal in normal brain comprising about 0.04 mg/g and essential for proper cellular function in normal physiological conditions (Cairo et al., 2006; Li et al., 2022a; Li et al., 2022b). Fe also is known to interact with other trace elements such as Mn and Cu since they share common transporters (e.g., DMT1) to cross the blood-brain barrier and enter the brain (Arredondo et al., 2006; Arredondo and Núñez, 2005). It is possible that elevated peripheral Mn and Cu levels may interfere with Fe binding to transporters resulting in suppressed brain Fe content despite elevated peripheral Fe levels (Fitsanakis et al., 2010). Moreover, once Fe enters the brain, it accumulates in a region-specific manner. For instance, MTL areas are known to contain considerably low Fe concentrations (Haacke et al., 2005; Rodrigue et al., 2011), whereas basal ganglia is a well-known area where Fe preferentially accumulates (Haacke et al., 2005). Thus, Fe deposition (exposure dose) and its meaning (or response) may vary in a region-specific manner, probably depending on speciation (Fe^3+^ vs Fe^2+^) and state (bound vs unbound). Therefore, since Fe has a complex relationship, as an external exposure and marker of cellular mechanism, its deposition may be beneficial for some brain areas such as MTL but detrimental in other regions such as basal ganglia, especially when the exposure level is relatively low as shown in the present study.

Second, we observed that higher whole blood Pb levels predicted higher entorhinal FA values, suggesting beneficial effects of Pb exposure on MTL DTI metrics. Blood Pb levels, however, were associated with lower executive function performance (*Phonemic Fluency* scores). These results appear contradictory at first. A subsequent mediation analysis suggested a direct link between higher whole blood Pb-lower executive function performance without significant involvement of the entorhinal FA metric. The negative impact of higher whole blood Pb on executive function is consistent with previous findings reporting lower cognitive performance in Pb-exposed subjects (Krieg Jr et al., 2005; Vlasak et al., 2019; Winker et al., 2006). The higher whole blood Pb-higher entorhinal FA association is intriguing and may partly reflect elevated Fe, not Pb, content in MTL structures that may have been induced by higher blood Pb. Prior animal studies reported increased brain Fe content following Pb exposure even though brain Pb content remained below the detection limit (Li et al., 2023; Zhu et al., 2013). It is also possible that higher brain Fe contents may show an reactive processes (Wang et al., 2023) when responding to elevated peripheral Pb, which may indirectly have led to higher entorhinal FA values.

Third, higher whole blood Zn levels predicted better executive function performance (higher *Trail Making-B* score). This result is consistent with a recent finding suggesting neuroprotective roles of Zn exposure in a set of neural damage biomarkers in metal-exposed welders (Wu et al., 2023). Zn is a second most abundant trace metal in brain (e.g., 10 µg/g) after Fe and plays vital roles in normal brain functions (Hussain et al., 2022; Lee, 2018; Roohani et al., 2013) although it can also be neurotoxic when deficient or overdosed (Hussain et al., 2022; Prasad, 2017; Skalny et al., 2021). The current finding should, however, be interpreted with caution because our exposed subjects (welders) eventually performed worse than controls on this *Trail Making-B* task. Although Zn exposure may have played a beneficial role to some degree, the elevated levels of other multiple metals may have affected cognitive performance in detrimental ways. Zn also is known to interact with Cu absorption to the body system in which Cu exposure may inhibit Zn from being absorbed or vice versa (Arredondo et al., 2006; Soliman et al., 2019). Thus, although both peripheral Cu and Zn levels are elevated in the present study, Zn absorption may have been inhibited by elevated Cu levels and thereby keeping Zn homeostasis in the brain.

## Limitations and conclusions

Together, the current findings suggest future studies need to consider both short and long-term mixed metal exposure history along with exposure intensity, collective dynamics of metal mixtures, as well as separately analyzing selective brain regions and metrics for cognitive outcomes. Our study has several limitations. First, we found no significant direct (?) collective or interaction effects of blood metal mixtures that were assessed by BKMR. This may be due partly to small sample sizes. The significant intercorrelations among blood metals (e.g., Cu, Fe, Mn, and Zn), however, imply potential interactions among metal mixtures to some degree. Future validation research should, where possible, utilize larger cohorts with advanced statistical models to better elucidate the dynamics of metal mixtures.

Second, we utilized questionnaire-based exposure history and whole blood metal levels to assess metal exposure dose. Exposure assessments that gauge long-term metal exposure effects should be employed to better delineate paths from mixed metal exposure to brain structural and neuropsychological consequences. We also measured several whole blood metal levels to estimate exposure dose of multiple metals, but the effects observed may be due to an unmeasured metal or co-occurring other environmental factors. Future studies assessing additional or alternate metals may provide further insight into the actions of metal co-exposures, something of both scientific and public health relevance.

Third, the exact neuropathological substrates underlying DTI metrics are unclear. It is speculated, however, that they reflect microstructural disorganization (*e.g.*, cytoarchitectural degeneration or demyelination processes) (Basser and Pierpaoli, 2011). Future studies with state-of-the-art MRI methods such as multi-shell DTI that utilizes multiple b-values to minimize motion or other artifacts and accurately model more detailed cellular features (Jha et al., 2022; Pines et al., 2020) may help dissect these neural mechanisms.

Fourth, it is important to note that although our exposed subjects had long work-related metal exposure history, they had, overall, relatively lower exposure levels compared to many other prior studies (*i.e.*, 11.0 *vs.* >14.2 μg/L for mean blood Mn; 22.7 *vs.* >30-50 μg/L for mean blood Pb) (Choi et al., 2007; Criswell et al., 2012; Jiang et al., 2008; Ko et al., 2017; Wu et al., 2022). This suggests other groups with relatively low, but chronic, exposure to the same metals as in our study may suffer similar sequalae, of great importance because chronic and low-level mixed metal exposures are ubiquitous.

## Data Availability

All data produced in the present study are available upon reasonable request to the authors.

## Funding

This work was supported by NIH grants R01 ES019672, R01 NS060722, U01s NS082151 and NS112008, the Hershey Medical Center General Clinical Research Center (National Center for Research Resources, UL1 TR002014), the Penn State College of Medicine Translational Brain Research Center, the PA Department of Health Tobacco CURE Funds, Basic Science Research Program through the National Research Foundation of Korea [2019R1G1A109957511; Ministry of Education (RS-2023-00247689)].

## Acknowledgements

We would like to thank all of the volunteers who participated in this study. In addition, we are indebted to many individuals who helped make this study possible, including: Melissa Santos, Tyler Corson, Lauren Deegan, and Susan Kocher for subject coordination, recruitment, blood sample handling, and data entry; Pam Susi and Pete Stafford of CPWR; Mark Garrett, John Clark, and Joe Jacoby of the International Brotherhood of Boilermakers; Fred Cosenza and all members of the Safety Committee for the Philadelphia Building and Construction Trades Council; Ed McGehean of the Steamfitters Local Union 420; Jim Stewart of the Operating Engineers; Sean Gerie of the Brotherhood of Maintenance of Way Employees Division Teamsters Rail Conference; and Terry Peck of Local 520 Plumbers, Pipefitters and HVAC.

## Conflicts of interest

The authors report no financial interests that relate to this research.

## Notes

### Competing Interest Statement

The authors have declared no competing interest.

### Author Declarations

The Internal Review Board (IRB) of Penn State Hershey gave ethical approval for this work.

### Summary of Updates

The background and interpretation have been revised, and new data analyses added.

